# Current Approaches to Evaluating Energy Requirements and Intake among Practicing Registered Dietitians

**DOI:** 10.1101/2025.07.21.25331863

**Authors:** Sarah A. Purcell, Tamara R. Cohen, Emilie K. Gosselin, Hilary Hildebrand, Vicky Drapeau, Shirin Panahi

## Abstract

**Background/Objectives:** Assessment of energy requirements and intake is central to the nutrition care process, yet current practices among registered dietitians (RDs) are not well characterized. This study examined how RDs assess energy requirements and intake, including perceived accuracy and resources, and differences by setting and experience.

**Methods:** A cross-sectional bilingual online survey was administered to RDs in Canada. The survey collected information on practice setting and experience, access to variables influencing energy requirements/intake, tool use, and opinions on accuracy and resource needs. Descriptive statistics and comparisons were made by practice setting (clinical, community, other) and years in practice (<5, 5–10, >10 years).

**Results:** 212 RDs completed the survey (62% clinical, 16% community, 22% other settings; 36% <5 years, 23% 5–10 years, 42% >10 years of practice). Participants rated importance of assessing energy requirements and energy intake as moderately high (6.8□±□2.2, 7.1□±□2.3 out of 10, respectively) and had regular access to variables needed to calculate energy requirements and intake (e.g., age, sex, weight, disease), although access to body composition, sleep, and stress was limited. Commonly-used tools included body weight-based equations and 24-hour recalls. Confidence was highest for delivering interventions and lowest for assessing intake (p < 0.001), especially among less experienced RDs (p = 0.002). Most respondents expressed interest in improved tools for assessing energy requirements (76%) and intake (74%).

**Conclusion:** Current RD practices vary, and access to key data is limited, underscoring the need for validated, accessible tools and training to support accurate energy assessment in dietetic care.

## Introduction

Registered dietitians (RDs) are regulated healthcare professionals who practice across a broad range of settings, including clinical environments (e.g., hospitals), research institutions, government agencies, private practice, community organizations, and corporate sectors. Within their scope of practice, RDs are responsible for delivering evidence-based nutrition interventions and medical nutrition therapy to diverse populations, supporting health and well-being, and offering services focused on food, nutrition, and related areas to promote public health across all age groups and socioeconomic backgrounds^1^. In light of the pivotal role that dietetic practice plays in patient-centred care and chronic disease prevention and management^2–6^, the demand for RDs in North America is projected to continue to grow at an above-average rate, underscoring their expanding importance in public health and clinical care^7,8^.

As the demand for RDs increases, understanding the common practices used by RDs across different practice settings is crucial for informing the development of evidence-based guidelines and the design of relevant research initiatives. Characterizing energy requirements and habitual energy intake is a key component to each stage of the nutrition care process: from assessing energy requirements/intake, diagnosing energy imbalances (e.g., excess or insufficient intake relative to requirements), providing energy balance-related interventions, to monitoring and evaluating changes in energy requirements or intake^9^. RDs have access to a diverse array of tools to assess various aspects of energy balance, including predicted or measured energy expenditure, mobile applications (‘apps’), food guides, and other resources; alternatively, some RDs may opt to focus on non-energy balance approaches, such as the Health at Every Size (HAES) model^10^. Despite the availability of these tools, there is limited understanding of the specific strategies RDs currently use to assess energy requirements or intake and how these practices vary across different contexts. A more comprehensive understanding of how RDs approach the assessment of energy requirements and intake could enhance current practices and inform future research aimed at refining nutrition assessment strategies that are both scientifically robust and directly applicable in dietetic practice settings. Thus, the main objective of this study was to evaluate available and currently utilized approaches for the assessment of energy requirements and intake used by Canadian RDs across various practice settings. We also characterized opinions regarding the accuracy of current approaches, the most commonly used resources for assessment, as well as differences according to practice setting and RD experience.

## Methods

### Study design

In this cross-sectional study, a bilingual survey was administered to RDs across Canada between June 20, 2024 and April 14, 2025. The study was conducted in two phases: 1) Development, refinement, and pilot testing of the survey, and 2) Data collection. Ethical approval was obtained from the University of British Columbia Behavioral Research Ethics Board (H22-01604). Given that the study involved only survey participation, informed consent was obtained through a cover letter containing the same information as a formal consent form, in accordance with institutional guidelines.

### Participants

Participant eligibility criteria were as follows: 1) valid registration as a RD in Canada, verified through provincial registration databases, 2) a minimum of 15 hours per week of direct patient or client interaction, or involvement in patient or client care planning, within their current practice setting for at least 3 months, and 3) proficiency in speaking and reading English or French. Only surveys with a completion rate of 75% or higher were included in the present analyses to minimize bias as those with lower completion rates often lacked responses to key questions. Participants were recruited through advertisements by Dietitians of Canada, the Ordre professionnel des diététistes du Québec, social media platforms (e.g., “Dietitians Support Group” and “Registered Dietitians and Future RDs” on Facebook), and word of mouth.

### Survey design and implementation

The initial version of the survey was developed by the research team based on previous literature, North American dietetic practice guidelines, and core concepts of energy balance. The draft survey was reviewed for clarity and relevance by two RDs. It was pilot tested between February 20, 2023 to May 3, 2023 and feedback from the pilot led to refinements in eligibility criteria, bot/validity screening, and clarification of several questions. The final version of the survey consisted of 44 to 73 items, depending on response patterns, as some questions in sections 3, 4, and 5 were conditionally displayed based on previous answers (**Supplementary Material**). Question formats included single response questions, multiple response questions, 5- or 10-point Likert scales, yes/no questions, and open-ended responses. The survey was organized into five sections: 1) Dietetic practice setting and history (5 questions; all single response), 2) General information about energy requirements/intake (6 questions; 4 single response, 2 slider scales), 3) Access and use of factors that may affect energy requirements (two tables with 30 items total, rated using 5-point Likert-type frequency and usage scales), 4) Tools for energy requirements, energy intake, and interventions (three tables with 29 items total, including up to 21 additional questions), and 5) Opinions on accuracy and helpful resources (up to 8 single response, multiple choice and open-ended responses). Free-text responses describing the populations served by participating RDs were categorized by a member of the research team (EKG) using a structured coding approach, with oversight and validation by a second investigator (SAP). A bilingual study co-ordinator translated the English version of the survey into French while a bilingual nutrition researcher (SP) back-translated the translated survey into English. The original survey and the back-translated version were compared by the two nutrition researchers (SP, VD) to identify any discrepancies, shifts in meaning, or ambiguities. Any inconsistencies were analyzed, and adjustments were made to the French version as needed. Free-text French responses were translated by a bilingual researcher (EKG). To ensure the sample reflected the national population distribution, the number of responses from each province was electronically capped based on the province’s population relative to the overall Canadian population. All data was collected using Qualtrics XM (Provo, UT, USA).

### Statistical analyses

As this was a descriptive study with no prior similar research to inform sample size calculations, determining an optimal sample size was neither appropriate nor feasible. An upper limit of 300 completed surveys was selected based on an estimated number of responses that could be reasonably obtained during the collection period. The final sample size was similar to previous surveys among RDs^10,11^.

All data were analyzed using SPSS (version 26; IBM, Armonk, NY, USA). Descriptive statistics are reported as mean ± standard deviation (SD) for normally distributed variables, median and interquartile range (IQR) for non-normally distributed variables, or frequencies and percentages for categorical variables. Normality was assessed using the Shapiro–Wilk test. Unless otherwise stated, p < 0.05 was considered statistically significant.

Between-group comparisons were conducted based on years of professional practice (<5 years, 5–10 years, >10 years; collapsed into three categories) and primary practice setting (clinical, community, or ‘other’; the latter collapsed due to small subgroup sizes). For continuous or ordinal data, Wilcoxon signed-rank tests were used to compare data from two independent groups, and Kruskal–Wallis tests with post hoc pairwise Mann–Whitney U tests (with Bonferroni correction, i.e., p = 0.017) were used to compare data across three independent groups. Subgroup comparisons for tool access and use were not performed due to low response frequencies in certain categories, which violated assumptions for Chi-square and non-parametric alternatives. Within-participant differences in responses were evaluated using Friedman’s test with post hoc Wilcoxon signed-rank tests for repeated-measures continuous variables (e.g., confidence ratings), and Cochran’s Q test with post hoc McNemar tests for paired categorical variables (e.g., frequency of seeking literature). McNemar–Bowker test of symmetry determined within-person differences in interest across multiple tools for energy requirements and intake (with three response levels: “yes,” “maybe,” “no”).

## Results

### Participants

A total of 336 initial survey responses were collected. Of these, 102 responses were excluded based on <75% completion of the survey and 22 individuals did not provide a valid RD registration number; thus, 212 responses were included in the final analysis.

Participants were employed across various dietetic settings, with the largest number of RDs working in clinical settings (61.9%), followed by community (16.2%) and private practice settings (11%), **Table 1**. The most common populations participants worked with were older adults (24%), individuals with diabetes (23.6%), or general populations/those in primary care (20.8%). Geographically, most participants were located in Ontario (39.2%), Quebec (19.3%), and British Columbia (16%). In terms of experience, most RDs had been practicing for 10-30 years (37.3%) and the most common length of time at the current place of employment was 1-5 years (40.1%).

**Table 1.** Characteristics of participants.

|  | **Category** | **n (%)** |
| --- | --- | --- |
| **Province/territory** | Ontario | 83 (39.2) |
|  | Quebec | 41 (19.3) |
|  | British Columbia | 34 (16.0) |
|  | Alberta | 18 (8.5) |
|  | Saskatchewan | 11 (5.2) |
|  | Manitoba | 6 (2.8) |
|  | New Brunswick | 6 (2.8) |
|  | Nova Scotia | 6 (2.8) |
|  | Prince Edward Island | 3 (1.4) |
|  | Northwest Territories | 2 (0.9) |
|  | Newfoundland | 1 (0.5) |
|  | Yukon | 1 (0.5) |
| **Work setting** | Clinical | 130 (61.9) |
|  | Community | 34 (16.2) |
|  | Private practice | 23 (11.0) |
|  | Other (free text responses) | 16 (7.6) |
|  | Corporate | 5 (1.9) |
|  | Food Service | 1 (0.5) |
|  | Research | 2 (1.0) |
|  | Missing | 2 (0.9) |
| **Population\*** | Older adults | 50 (23.6) |
|  | Diabetes | 50 (23.6) |
|  | All populations/primary care | 44 (20.8) |
|  | Other | 41 (19.3) |
|  | Bariatric/weight loss/obesity | 23 (10.8) |
|  | Cancer | 20 (9.4) |
|  | Cardiac | 19 (9.0) |
|  | General healthy | 19 (9.0) |
|  | Pediatrics | 19 (9.0) |
|  | General chronic disease | 18 (8.5) |
|  | Acute care | 15 (7.1) |
|  | Gastrointestinal | 15 (7.1) |
|  | Renal | 11 (5.2) |
|  | Athletes | 7 (3.3) |
|  | Eating disorders | 7 (3.3) |
|  | Indigenous/Rural | 5 (2.4) |
|  | Mental health | 5 (2.4) |
|  | Prenatal | 5 (2.4) |
| **Years as a registered dietitian** | 6-12 months | 5 (2.4) |
|  | 1-5 years | 71 (33.5) |
|  | 5-10 years | 48 (22.6) |
|  | 10-30 years | 79 (37.3) |
|  | >30 years | 9 (4.2) |
| **Duration at current workplace** | 3-6 months | 17 (8.0) |
|  | 6-12 months | 20 (9.4) |
|  | 1-5 years | 85 (40.1) |
|  | 5-10 years | 37 (17.5) |
|  | 10-30 years | 47 (22.2) |
|  | >30 years | 6 (2.8) |

N=212 participants. No valid responses were received from individuals in Nunavut.
\*total number does not equal 212, as participants could list multiple populations.

### General questions about energy requirements and habitual energy intake

Participants rated the importance of assessing energy requirements and energy intake (assessed by 10-point Likert scale) as moderately high, with mean scores of 6.8L±L2.2 and 7.1L±L2.3, respectively. There were no differences in the importance of assessing energy requirements (H(2) = 3.34, p = 0.188) or intake (H(2) = 5.13, p = 0.077) according to number of years of practice or in the importance of assessing energy intake across practice settings (H(2) = 4.48, p = 0.106). However, there was a significant difference in the importance of assessing energy requirements (H(2) = 14.85, p < 0.001), with RDs in clinical settings rating this as more important than those in community (p < 0.001) and other settings (p = 0.024).

Approximately 70% of respondents reported assessing some aspect of energy requirements or energy intake at least occasionally, **Figure 1**. In contrast, only a small proportion of participants reported discussing concepts related to energy balance in 80% or more of their patient/client interactions. There were no differences in the frequency of discussing energy balance (H(2) = 1.41, p = 0.495) or assessing energy requirements (H(2) = 0.88, p = 0.644) or energy intake (H(2) = 1.14, p = 0.564) according to years of dietetic practice. There were also no differences in how frequently RDs reported discussing energy balance (H(2) = 1.72, p = 0.424) or assessing energy intake (H(2) = 2.65, p = 0.266) across practice settings. Frequency of energy requirements assessment differed by practice setting (H(2) = 14.78, p < .001), with clinical RDs reporting higher frequency than those in community settings (p < .001) and a non-significant trend compared to other settings after Bonferroni correction (p = 0.023).

**Figure 1.**
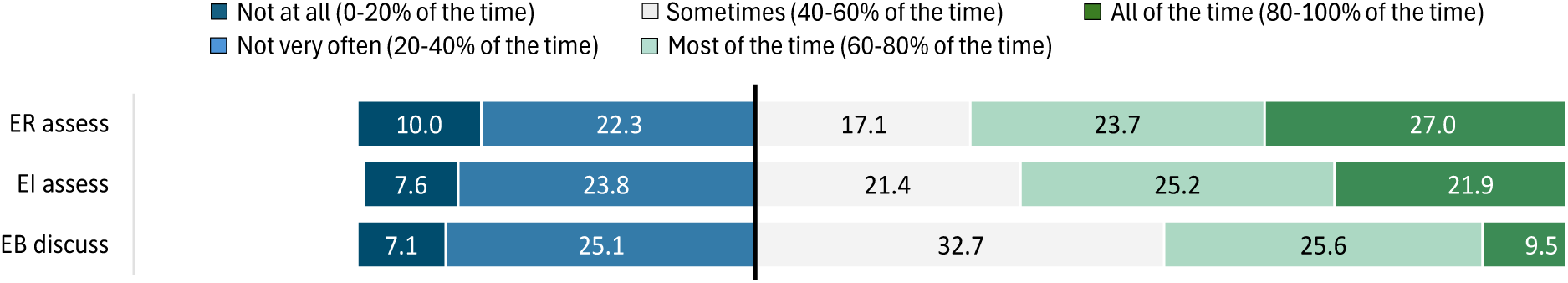
Assessment of energy requirements (ER) or energy intake (EI) or discussion of energy balance (EB) frequency by registered dietitians.

### Access and use of factors affecting energy requirements

The majority of participants (>90%) reported having consistent access to patient/client age and sex for energy requirements assessment at all times (**Figure 2a**); these variables were also the commonly used as illustrated in **Figure 2b**. Disease status, body weight/body mass index (BMI), weight history, and physical activity were among the most frequently accessed and used factors in determining energy requirements. Access to other sociodemographic variables was relatively evenly distributed across the frequency categories. Many of these variables were among the least frequently used to determine energy requirements. Over 50% of participants indicated they had access to information on sleep and stress at least sometimes, but fewer reported using this information to estimate energy requirements. Only ∼20-25% of participants indicated they had access to or used body composition information at least sometimes.

**Figure 2.**
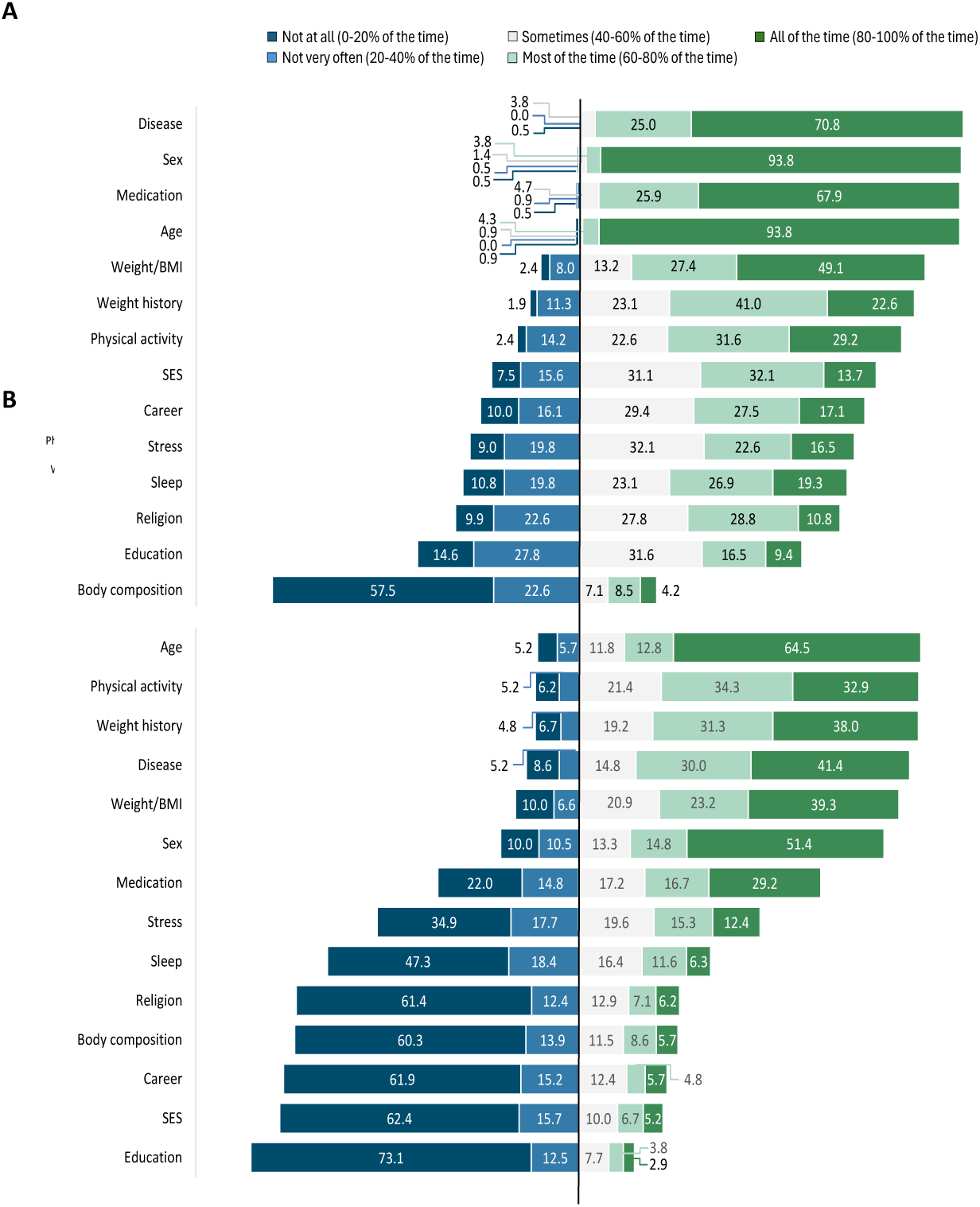
Frequency registered dietitians have access to (A) or use (B) factors that may impact energy requirements. BMI: body mass index; SES: socioeconomic status.

### Tools/approaches for energy requirements, energy intake, and nutrition interventions

The use of tools for estimating energy requirements varied widely (**Figure 3a**). Indirect calorimetry was rarely used, with over 90% of respondents reporting that they never use it. Similarly, apps and consideration of the thermic effect of food in energy requirements calculations were used infrequently. Body-weight based equations and ideal/adjusted body weight were frequently used, with over half of RDs indicating they used these approaches most or all of the time. The use of the Dietary Reference Intakes (DRIs), disease factors, resting metabolic rate-based equations, and physical activity coefficients varied, with approximately half or more reporting the use of these factors at least sometimes.

**Figure 3.**
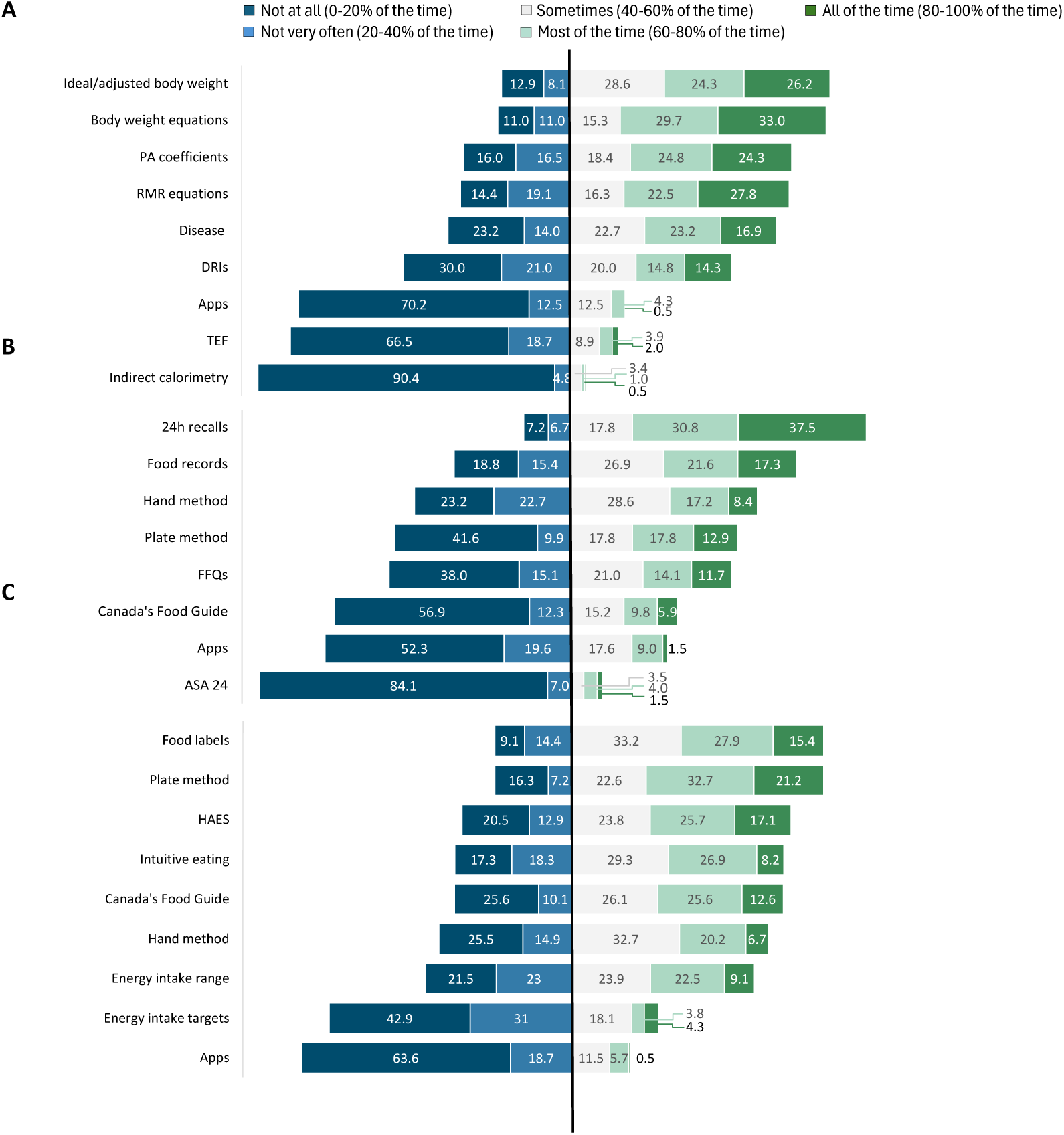
Frequency of tools and approaches for assessing energy requirements (A), energy intake (B), or implementing nutrition interventions (C) among registered dietitians. Apps: Applications (mobile); ASA24: Automated Self-Administered 24-hour Dietary Assessment Tool; DRIs: Dietary reference Intakes; FFQs: Food frequency questionnaires; HAES: Health at Every Size; PA: physical activity coefficient; TEF: Thermic effect of food.

Dietary assessment platforms such as the Automated Self-Administered 24-hour Dietary Assessment Tool^12^, mobile diet applications (apps), and Canada’s Food Guide were rarely used to assess energy intake, with over half of participants indicating that they never used them (**Figure 3b**). More traditional methods such as 24-hour dietary recalls and food records (with no specific number of days) were more commonly utilized; 37.5% and 17.3% of respondents reported using these methods all of the time, respectively, and another ∼22–31% used them most of the time. The hand method, plate method and FFQs were moderately used.

Diet apps and specific energy intake targets were the least frequently used strategies for nutrition interventions, with over 60% and 40% of respondents, respectively, indicating they did not use these tools or approaches (**Figure 3c**). In contrast, practical strategies such as the plate method (e.g., visually dividing the plate into portions for vegetables, protein, and grains) and use of food labels were more common; these were used all or most of the time by 53.9% and 43.3% of respondents, respectively. Approaches such as intuitive eating, HAES, and Canada’s Food Guide were used with moderate frequency, while the hand method and energy intake ranges were less consistently applied.

### Opinions on tools and resources

Most RDs were uncertain about the accuracy of the tools they use to estimate energy requirements or energy intake; 46.1% responded “maybe” when asked about the accuracy of energy requirements tools, while 54.9% responded “maybe” for energy intake tools. Fewer RDs believed the tools were accurate (22.8% for energy requirements, 19.1% for energy intake), and approximately one-third indicated they were not accurate (31.1% and 26.0%, respectively). Participants reported significantly greater confidence in providing effective nutrition advice or interventions (8.4 ± 1.1) compared to estimating energy requirements (7.6 ± 1.7), and both were rated higher than confidence in assessing energy intake (7.0 ± 1.7; all p < 0.001 for main and post hoc comparisons). Confidence in assessing energy intake differed by years of practice (H(2) = 6.31, p = 0.043); however, but post hoc comparisons were not significant after Bonferroni correction. Confidence in providing nutrition interventions also differed by experience level (H(2) = 12.53, p = 0.002), with lower confidence among RDs with <5 years compared to those with 5–10 years (p = 0.003) and >10 years (p = 0.002) of experience. Clinical RDs reported greater confidence in assessing energy requirements than those in community settings (H(2) = 6.94, p = 0.031), although this difference did not reach significance after Bonferroni correction (p = 0.021).

Participants were less likely to report seeking information on tools to estimate habitual energy intake (34.3%) compared to seeking literature on how to estimate energy requirements (84.7%; χ² = 98.1, p<0.001) and provide nutrition interventions (75.7%; χ² = 73.2, p<0.001) (Cochran’s Q = 146.7, p < 0.001). The most common sources of this information were research papers or literature reviews, dietetic-specific resources (e.g., Practice-based Evidence in Nutrition^13^), and conferences, webinars, or presentations (**Figure 4**).

**Figure 4.**
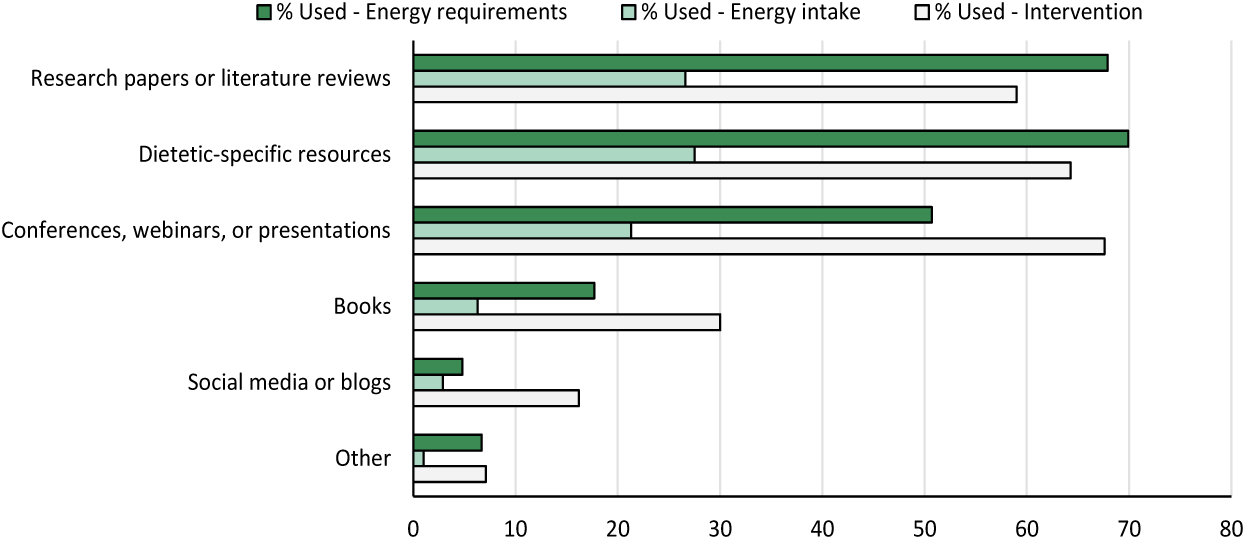
Sources of information registered dietitians use to explore energy requirements, energy intake, and nutrition interventions.

Most participants expressed interest in tools to improve assessment of energy requirements (76.0%) and intake (73.6%). Fewer were uncertain (14.4% and 17.8%, respectively), and less than 10% were not interested. Interest did not differ significantly between the two content areas (χ² = 1.72, p = 0.633). Dietetic-specific resources, apps or online calculators, body composition tools, and population-specific equations were among the most common tools that RDs expressed interest in (**Figure 5**). Among RDs currently using indirect calorimetry (n=18), the majority (61.1%) reported that it enhances their ability to provide effective nutrition care, while 16.7% disagreed and 22.2% were uncertain. In contrast, among those not using indirect calorimetry (n=194), 34.8% indicated that it would improve care, 39.8% indicated it would not, and 25.4% were unsure.

**Figure 5.**
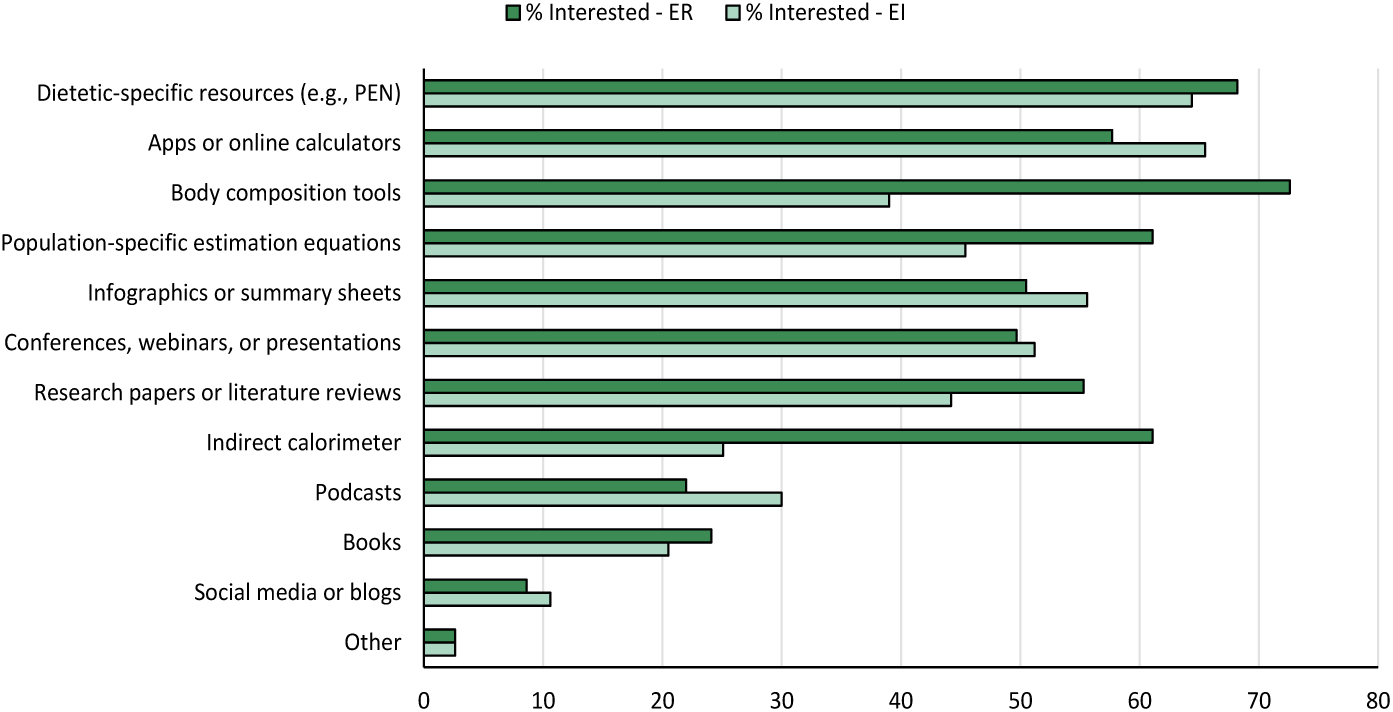
Resources or tools that registered dietitians indicate would be most helpful in estimating energy requirements or energy intake. PEN: Practice-based Evidence in Nutrition.

## Discussion

To our knowledge, this study is the first to examine current practices among RDs for assessing energy requirements and energy intake across diverse settings. RDs rated both energy requirements and intake assessment as important and reported frequent access to core variables such as age, sex, body weight, and disease status, while access to factors such as body composition, sleep, stress, and sociodemographic data was limited. Use of tools to estimate energy requirements and intake varied widely among RDs, with practical strategies (e.g., plate method, food labels) most commonly applied. Confidence was highest for providing nutrition interventions and lowest for assessing energy intake, particularly among less experienced RDs. The majority of participants expressed interest in improved tools for assessing both energy requirements and intake. These findings highlight the need for more accessible, validated tools to support accurate energy assessment in ‘everyday’ dietetic practice.

Evaluation of energy requirements and intake is a critical component of the ‘assessment’ phase of the nutrition care process^9^ and therefore fundamental to evidence-based nutrition practice. This was reflected in our findings, where RDs rated both energy requirement and intake assessment as important, with most reporting that they assess these factors at least sometimes. Clinical RDs in particular rated energy requirements assessment as more important and reported using it more frequently. This is not unexpected, as chronic disease is a common reason for dietetic referral^14,15^, and disease-related factors—such as inflammation, medication use, and altered hormones or body composition—can significantly affect energy requirements^16–22^. Moreover, the goals of care in clinical populations often extend beyond general health promotion to include disease management, prevention of malnutrition, and tailored nutrition support – which could require more precise estimation of energy requirements. These findings reinforce the importance of context-specific tools and guidance in supporting accurate energy assessments, particularly in clinical settings.

In this study, RDs most commonly reported having access to and using factors such as age, sex, disease status, body weight/BMI, weight history, and physical activity when estimating energy requirements. Approaches such as ideal or adjusted body weight and predictive equations were also frequently used. These findings are expected, given that age, body weight, and physical activity are well-established contributors to total energy expenditure and thus energy requirements^23,24^. While disease status can also influence energy expenditure, the direction and magnitude of its impact are highly variable and context-specific^25^. Given that population-specific energy requirements equations were among the most identified tools of interest by RDs, further research is warranted to develop and implement such tailored tools.

Measured body composition and resting metabolic rate via indirect calorimetry were rarely used for energy requirements assessments. This finding is consistent with prior research that identifies barriers to body composition assessment in clinical practice, including a lack of awareness among healthcare professionals, insufficient integration into healthcare workflows, limited access to equipment or imaging, time constraints, and uncertainty about when and with whom assessments should be conducted^26–29^. Although no studies to date have systematically examined barriers to indirect calorimetry across different dietetic practice settings, limited research in intensive care units suggests that lack of access to validated devices remains a key barrier^30^. The infrequent use of indirect calorimetry represents a gap between current practice and clinical guidelines, which recommend its use in several contexts—including critical care and, increasingly, in outpatient settings such as obesity management^2,31–33^. Emerging evidence suggests that incorporating measured resting metabolic rate may enhance the accuracy of weight management strategies, support monitoring of adherence to nutritional interventions, and contribute to the prediction of disease trajectories^35–37^. The perceived clinical utility of indirect calorimetry was evident in our sample, with the majority of RDs indicating that it would or could be a valuable tool in practice. However, its feasibility, accuracy in improving energy requirements estimations, and potential impact on patient outcomes outside of intensive care units remain unclear and warrant further investigation.

Given the central role of energy intake in managing nutritional status and health outcomes, we also investigated how RDs assess intake and implement nutrition-related interventions. Although self-reported dietary intake methods are known to underestimate true intake^38^, traditional approaches such as 24-hour dietary recalls and food records were commonly used by RDs in this study to assess energy intake. Similarly, practical strategies such as reading food labels and using the plate method were frequently employed in nutrition interventions. These preferences likely reflect the accessibility, low cost, and ease of implementation of these tools.

Non-weight-focused approaches, including HAES and intuitive eating, were used with moderate frequency, potentially reflecting their relevance to specific populations and practice contexts. This aligns with existing literature indicating that such frameworks may promote improvements in dietary quality, eating behaviours, weight stability, and psychological well-being^39,40^, with mixed evidence on outcomes such as weight loss, cardiovascular health, or physical activity^39,41^. Interestingly, the use of Canada’s Food Guide for nutrition intervention was reported at low to moderate levels in our sample, contrasting with earlier findings where Canada’s Food Guide was among the most frequently recommended strategies for patients with overweight or obesity^42^. This may reflect changes in clinical practice, evolving evidence, or differing perceptions of the guide’s applicability for individualized care. RDs also reported infrequent use of the hand-based portion estimation method, which is somewhat unexpected given its potential utility in populations with low literacy or numeracy skills.

Despite increasing research interest in digital health among consumers^43^, apps were infrequently used for both energy intake assessment and intervention. This underutilization contrasts with growing evidence supporting the effectiveness of app-based interventions in improving dietary behaviours and related health outcomes^44^. Reported barriers such as app cost, usability challenges, technical issues, and user motivation may explain their limited adoption in practice^45^. Similarly, food composition databases, lack of local database support, and technological proficiency are barriers to recommending nutrition apps among healthcare professionals^46^. Other dietary self-monitoring tools that resemble the plate-based approach to eating, such as iCANPlate^47^, could help facilitate healthy eating practices, although more research in needed to understand how they would help RDs calculate energy requirements. Collectively, these findings indicate that while RDs primarily rely on accessible and familiar tools, further research is needed to determine whether and how other methods can support the implementation of emerging evidence-based approaches in routine nutrition care.

Most RDs expressed uncertainty regarding the accuracy of tools used to assess energy requirements and energy intake, and reported lower confidence in these areas compared to providing nutrition interventions—particularly among those with fewer years of professional experience. This aligns with previous research indicating that impostor syndrome is more prevalent among early-career RDs compared to those with more experience^48^, suggesting that feelings of impostorism decrease as RDs gain experience, observe positive patient/client outcomes, and develop greater expertise. Our findings also have implications for future research and tool development. When seeking guidance, RDs most commonly relied on evidence-based resources such as peer-reviewed literature, professional practice platforms, and educational events. This reliance on credible sources is encouraging and underscores the importance of ensuring future tools are scientifically validated and easily integrated into such platforms.

Strengths of this study include the inclusion of both English- and French-speaking RDs and broad representation across Canadian provinces and various practice settings. However, several limitations should be noted. Recruitment through professional networks, listservs, and social media may have introduced sampling bias, and the sample size is small relative to the national population of RDs. Additionally, the use of self-reported data introduces the potential for measurement error due to reporting bias. However, we were able to recruit a variety of practice areas-beyond clinical nutrition-which helps increase the generalizability of our findings. Finally, to better understand the lower confidence reported by early-career RDs, research is needed to examine the extent and nature of content related to energy metabolism and energy requirements within Canadian dietetic curricula. This would help determine whether foundational knowledge is being adequately addressed during academic training or primarily acquired during practice education, and whether current entry-to-practice competencies are being effectively met across both settings.

Assessing energy requirements and intake is crucial for practicing dietitians to deliver effective, evidence-based nutrition care. Accurate energy assessment ensures personalized dietary recommendations that align with individual needs, supporting optimal health outcomes, disease management, and prevention of malnutrition or obesity. Inaccurate assessments can lead to suboptimal interventions, compromising patient health and treatment efficacy. For dietitians, understanding energy balance is foundational to tailoring interventions for diverse populations, from clinical patients to athletes, across various practice settings. The novelty of the study lies in its exploration of gaps in access to and confidence with energy assessment tools among dietitians, revealing practical barriers to integrating advanced methods like body composition tools, indirect calorimetry, and dietary apps. Given the broad interest in improved tools for assessing energy requirements and intake, future efforts should prioritize the development of accessible, evidence-based methods that enhance accuracy, usability, and clinical relevance. Such tools will be essential for advancing the quality and consistency of evidence-informed nutrition care. Importantly, these efforts must be supported by comprehensive training, particularly for new and early-career dietitians. Ensuring that emerging professionals are equipped with the skills and confidence to use advanced assessment methods will be essential for strengthening the quality and consistency of evidence-informed nutrition care.

## Data Availability

All data produced in the present study are available upon reasonable request to the authors.

## Data Availability Statement

Due to ethical restrictions related to participant privacy, the data that support the findings of this study are not publicly available. However, anonymized data may be made available from the corresponding author upon reasonable request.

## Acknowledgments

We thank all participants for their participation in the study.

## Author contributions

SAP, TRC, and HH contributed to the study design and survey development. SAP, HH, VD, and SP contributed to data collection. SAP and EKG contributed to data cleaning and analyses. SAP wrote the initial manuscript draft. All authors contributed to revising the manuscript and have read and approved the final version.

## Ethical Approval

This study was approved by the University of British Columbia Behavioral Research Ethics Board (H22-01604). Given that the study involved only survey participation, informed consent was obtained through a cover letter containing the same information as a formal consent form, in accordance with institutional guidelines.

