## Supplementary material for "Current Approaches to Evaluating Energy Requirements and Intake among Practicing Registered Dietitians"

### Energy Intake Assessment Practices of Registered Dietitians in Canada

#### Part 1: Dietetic practice setting and history

1. What type of dietetic setting do you work in?
    - Community
    - Research
    - Clinical (e.g., hospital, specialist outpatient)
    - Private Practice
    - Corporate
    - Food Service
    - Other (please specify):

2. What population do you work most closely with? (e.g., people with cancer, people with diabetes, athletes, ICU inpatients, etc.)

3. Are patients counselled?
    - Yes
    - No

4. How long have you worked as a RD in **total**? If you took a career break or parental or sick leave, please estimate the total time you have actively worked as a RD without the time away from active practice. In other words, please subtract any leave from the total duration you have worked as a RD.
    - <3 months
    - 3-6 months
    - 6-12 months
    - 1 to 5 years
    - 5 to 10 years
    - 10 to 30 years
    - >30 years

5. What province/ territory do you work in?
    - Alberta
    - British Columbia
    - Manitoba
    - New Brunswick
    - Newfoundland and Labrador
    - Northwest Territories
    - Nova Scotia
    - Nunavut
    - Ontario
    - Prince Edward Island
    - Quebec
    - Saskatchewan
    - Yukon

#### Part 2: General questions about energy intake

In the following questions, '**energy requirements**' is the estimated dietary energy ('calories') intake a person needs to maintain energy balance and meet their nutrition or body weight goal (e.g., weight loss, maintenance, gain). '**Habitual energy intake**' is the energy a person consumes on a regular basis. The following questions ask about your interactions with your patient/clients or how you assess requirements or prescribe recommendations; they are not referring to your interactions with other RDs or any outside research you may do.

1. How often do you discuss any concept related to energy balance (e.g., energy consumed, energy expenditure)?
    - Not at all *(0-20% of the time)*
    - Not very often *(~20-40% of the time)*
    - Sometimes *(~40-60% of the time)*
    - Most of the time *(~60-80% of the time)*
    - All of the time *(~80-100% of the time)*

2. In your practice, how often do you assess a patient/client's **energy requirements** by any means as part of the nutrition care process?
    - Not at all *(0-20% of the time)*
    - Not very often *(~20-40% of the time)*
    - Sometimes *(~40-60% of the time*
    - Most of the time *(~60-80% of the time)*
    - All of the time *(~80-100% of the time)*

3. On a scale of 1 - 10 with 1 being not important at all and 10 being very important, how important is assessing a patient/client's **energy requirements**?
    *[slider scale from 1 to 10 in REDCap]*
    3a. Why do you think assessing a patient/client's **energy requirements** is/is not important? *[typed response]*

4. In your practice, how often do you assess a patient/client's **habitual energy intake** as part of the nutrition care process?
    - Not at all *(0-20% of the time)*
    - Not very often *(~20-40% of the time)*
    - Sometimes *(~40-60% of the time)*
    - Most of the time *(~60-80% of the time)*
    - All of the time *(~80-100% of the time)*

5. On a scale of 1 - 10 with 1 being not important at all and 10 being very important, how important is assessing a patient/client's **habitual energy intake**?
    *[slider scale from 1 to 10 in REDCap]*
    5a. Why do you think assessing a patient/client's **habitual energy intake** is/is not important? *[typed response]*

6. Are there any words or phrases related to energy intake or energy balance that you avoid using in practice (e.g., "I often avoid the term 'calories' with eating disorder patients")?
    - Yes
    - No
    - *[If yes, what words or phases do you avoid?]:*
    -

**Part 3: Access to factors affecting energy intake assessment**

Note the following questions ask about the **availability and your use** of factors that may impact **energy requirements** at any part of the nutrition care process (i.e., assessment, diagnosis, intervention, monitoring/evaluation). This can include things you or the dietetic team measure specifically for dietetic practice or things already in a patient/client's medical record or intake forms.

For energy requirement assessment, how often do you **have access to** a patient/client's:

| | **Not at all** *(~0-20% of the time)* | **Not very often** *(~20-40% of the time)* | **Sometimes** *(~40-60% of the time)* | **Most of the time** *(~60-80% of the time)* | **All of the time** *(~80-100% of the time)* |
|---|---|---|---|---|---|
| Age | | | | | |
| Sex | | | | | |
| Socioeconomic status | | | | | |
| Career | | | | | |
| Education level | | | | | |
| Religious/cultural practices related to nutrition | | | | | |
| Body weight or body mass index (BMI) | | | | | |
| Weight history | | | | | |
| Body composition (e.g., bioelectrical impedance analysis [BIA]) or any other anthropometrics to estimate body composition (e.g., waist circumference, calf circumference) | | | | | |
| Physical activity or exercise habits | | | | | |
| Sleep habits | | | | | |
| Chronic diseases | | | | | |
| Medication/supplement use | | | | | |
| Stress levels | | | | | |
| Other (please specify): | | | | | |

For energy requirement assessment, how often do you **use** the following?

| | **Not at all** *(~0-20% of the time)* | **Not very often** *(~20-40% of the time)* | **Sometimes** *(~40-60% of the time)* | **Most of the time** *(~60-80% of the time)* | **All of the time** *(~80-100% of the time)* |
|---|---|---|---|---|---|
| Age | | | | | |
| Sex | | | | | |
| Socioeconomic status | | | | | |
| Career | | | | | |
| Education level | | | | | |
| Religious/cultural practices related to nutrition | | | | | |
| Body weight or body mass index (BMI) | | | | | |

| |
|---|
| Weight history |
| Body composition (e.g., bioelectrical impedance analysis [BIA]) or any other anthropometrics to estimate body composition (e.g., waist circumference, calf circumference) |
| Physical activity or exercise habits |
| Sleep habits |
| Chronic diseases |
| Medication/supplement use |
| Stress levels |
| Other (please specify): |

**Part 4: Tools for energy requirements, energy intake ,and interventions**
Note the following questions ask about the availability and your use of tools for **energy requirements** (part 4a), **habitual energy intake** (part 4b), and **nutrition intervention** (part 4c).

**4a: Energy requirements**
1. On a scale of 1 to 10, with 1 being not confident at all to 10 being extremely confident, how confident are you in your knowledge on your ability to estimate a patient/client's **energy requirements**? *[slider scale from 1 to 10 in REDCap]*

2. Have you ever explored literature on what impacts **energy requirements** or how to estimate **energy requirements**?

- Yes
- No

2a. (if yes): What was the source of this information?

- Research papers or literature reviews
- Dietetic-specific resources (e.g., Practice-based Evidence in Nutrition)
- Conferences, webinars, or presentations
- Books
- Social media or blogs
- Other (please specify): ____________________

How often do you use the following to assess **energy requirements?**

| | **Not at all** *(~0-20% of the time)* | **Not very often** *(~20-40% of the time)* | **Sometimes** *(~40-60% of the time)* | **Most of the time** *(~60-80% of the time)* | **All of the time** *(~80-100% of the time)* |
|---|---|---|---|---|---|
| Equations for resting/basal metabolic rate | | | | | |
| Adjustment or coefficient for physical activity | | | | | |
| Adjustment or coefficient for the thermic effect of food | | | | | |
| Adjustment or coefficient for disease-specific factors | | | | | |

| |
|---|
| Ideal or adjusted body weight |
| Dietary reference intake equations |
| Equations based on kcal/kg (e.g., 30 kcal/kg/day) |
| Indirect calorimetry (e.g., 'metabolic carts' like MedGem, Q-NRG) |
| Apps on a mobile phone (e.g., MyFitnessPal) |
| Other (please specify): |

*If a participant indicates they use equations for resting/basal metabolic rate:*
3a. What equation(s) do you use? (e.g., Harris-Benedict, Mifflin St-Jeor, Dietary Reference Intake, etc.)

3b. Do you use the same or different equations for all patients/clients?

- Same
- Different
    - *[if different, please explain]:*

*If a participant indicates they adjust for physical activity:*
4a. What activity factors or coefficients do you use? (e.g., resting metabolic rate x 1.3, physical activity coefficient of 1.12 in the DRI equation, etc.)

4b. Do you use the same or different activity factors or coefficients for all patients/clients?

- Same
- Different
    - *[if different, please explain]:*

*If a participant indicates they adjust for disease-specific factors:*
5a. What disease-specific adjustments do you use? (e.g., resting metabolic rate x 1.3 for people with cancer)

5b. Do you use the same or different adjustments for all patients/clients with the same disease/condition?

- Same
- Different
    - *[if different, please explain]:*

*If a participant indicates they use an indirect calorimeter:*
6a. What brand/model of indirect calorimeter do you use?

6b. Do you think the indirect calorimeter you use helps you provide better nutrition care?

- Yes
- No
- Maybe

Explain: ___________________

*If a participant indicates they do not use an indirect calorimeter:*
6c. If you were to have access to an indirect calorimeter for assessing energy requirements, do you think it would be helpful for your practice?

- Yes

- No
- Maybe

Explain:______________________

*If a participant indicates they use apps:*
7. What apps do you find most helpful in assessing energy requirements?

**4b. Habitual energy intake**
8. On a scale of 1 to 10, with 1 being not confident at all to 10 being extremely confident, how confident are you in your knowledge on your ability to estimate a patient/client's **habitual energy intake**? *[slider scale from 1 to 10 in REDCap]*

9. Have you ever explored literature on the **best tools to estimate habitual energy intake**?
- Yes
- No

9a. (if yes): What was the source of this information?
- Research papers or literature reviews
- Dietetic-specific resources (e.g., Practice-based Evidence in Nutrition)
- Conferences, webinars, or presentations
- Books
- Social media or blogs
- Other (please specify): ___________________

How often do you use the following to assess **habitual energy intake?**

| | **Not at all** *(~0-20% of the time)* | **Not very often** *(~20-40% of the time)* | **Sometimes** *(~40-60% of the time)* | **Most of the time** *(~60-80% of the time)* | **All of the time** *(~80-100% of the time)* |
|---|---|---|---|---|---|
| Food frequency questionnaires (FFQs) | | | | | |
| 24-hour recall | | | | | |
| Automated Self-Administered Recall System (ASA24) | | | | | |
| Multiple-day food records | | | | | |
| 2019 Canadian Food Guide | | | | | |
| Plate method (e.g., My Plate) | | | | | |
| Apps on a phone (e.g., MyFitnessPal) | | | | | |
| "Hand" method (e.g., clenched fist is ~1 cup) | | | | | |
| Other (please specify) | | | | | |

*If a participant indicates they use apps:*
10. What apps do you find most helpful in assessing **habitual energy intake**?

**4c: Nutrition intervention**

11. On a scale of 1 to 10, with 1 being not confident at all to 10 being extremely confident, how confident are you in your knowledge on your ability to **providing effective nutrition advice or interventions**? *[slider scale from 1 to 10 in REDCap]*

12. Have you ever explored literature on the **best methods for providing effective nutrition advice or interventions**?

- Yes
- No

12a. (if yes): What was the source of this information?

- Research papers or literature reviews
- Dietetic-specific resources (e.g., Practice-based Evidence in Nutrition)
- Conferences, webinars, or presentations
- Books
- Social media or blogs
- Other (please specify): ___________________

How often do you use the following for providing **nutrition advice or interventions?**

| | **Not at all** *(~0-20% of the time)* | **Not very often** *(~20-40% of the time)* | **Sometimes** *(~40-60% of the time)* | **Most of the time** *(~60-80% of the time)* | **All of the time** *(~80-100% of the time)* |
|---|---|---|---|---|---|
| Specific energy intake targets (e.g., 2050 kcal/day) | | | | | |
| A range of energy intake targets (e.g., 2000-2500 kcal/day) | | | | | |
| Health at Every Size approach | | | | | |
| Intuitive eating | | | | | |
| 2019 Canada's Food Guide | | | | | |
| Plate method (e.g., My Plate) | | | | | |
| Apps on a phone (e.g., MyFitnessPal) | | | | | |
| "Hand" method (e.g., clenched fist is ~1 cup) | | | | | |
| Food label interpretation | | | | | |
| Other (please specify) | | | | | |

*If a participant indicates they use apps:*

13. What apps do you find most helpful in providing nutrition advice or interventions?

**<u>Part 5: (last part): Open-ended responses</u>**

1. Do you think the tools RDs use to assess **energy requirements** are accurate?
   - Yes
   - No
   - Maybe

     Explain:______________________

2. Do you think the tools RDs use to assess **habitual energy intake** are accurate?
   - Yes
   - No
   - Maybe
   
   Explain:______________________
3. If you use tools to assess energy requirements or habitual energy intake, which ones are most useful to you? What makes them most useful?

4. Would you be interested in resources or tools to help you better assess **energy requirements**?
   - Yes
   - No
   - Maybe

4a. (if yes or maybe): What types of resources or tools would be most helpful to you?
- Research papers or literature reviews
- Dietetic-specific resources (e.g., Practice-based Evidence in Nutrition)
- Conferences, webinars, or presentations
- Podcasts
- Books
- Social media or blogs
- Infographics or summary sheets
- Apps or online calculators
- Indirect calorimeter
- Body composition tools
- Population-specific estimation equations
- Other (please specify): ___________________

5. Would you be interested in resources or tools to help you better assess **habitual energy intake**?
   - Yes
   - No
   - Maybe

5a. (if yes or maybe): What types of resources or tools would be most helpful to you?
- Research papers or literature reviews
- Dietetic-specific resources (e.g., Practice-based Evidence in Nutrition)
- Conferences, webinars, or presentations
- Podcasts
- Books
- Social media or blogs
- Infographics or summary sheets
- Educational sheets for patient/clients
- Apps or online tools
- Population-specific factors to adjust for under- or over-reporting
- Other (please specify): ___________________

6. Is there anything else you would like to add about how you currently assess energy intake, what you would like to do in the future, or how research could help inform dietetic practice around energy balance?
